# The Conundrum of Giglio Island: unraveling the dynamics of an apparent resistance to COVID-19 – A descriptive study

**DOI:** 10.1101/2021.01.08.20248948

**Authors:** Antonio Bognanni, Armando Schiaffino, Fulvia Pimpinelli, Sara Donzelli, Ilaria Celesti, Sabrina Strano, Elena Solari, Giorgia Schiaffino, Gabriele Solari, Domenico Solari, Serena Delbue, Marta Rigoni, Giandomenico Nollo, Greta E. Muti, Giovanna E.U. Muti Schünemann, Holger J Schünemann, Giovanni Blandino, Aldo Morrone, Paola Muti

## Abstract

**Objectives:** Despite an extensive risk of exposure to COVID-19, the residents of Giglio Island, Italy, did not develop any symptom of SARS-CoV-2. The present study aims to characterize the nature of exposure and to describe the local population dynamics underlying its apparent resistance to COVID-19.

**Methods:** We conducted seroprevalence screening from April 29 to May 3, 2020 across the three main settlements on the island. We invited the adult resident population, present on the island to undergo testing by rapid serologic assay and to provide a sample of saliva for molecular validation. We monitored the participation through the official municipality residents’ list. Serologic testing was performed using a COVID-19 IgG/IgM rapid test while molecular analyses were carried out by Allplex 2019-nCoV Assay (Seegene).

**Results:** A total of 634 residents out of 748 (84.8%) present at the time, and 89 non-residents underwent serological testing. 364 males and 359 females with a median age of 58.5 years. The serological screening identified one positive, asymptomatic subject. The Nucleic Acid Amplification Tests did not yield any positive result.

**Conclusion:** Despite extensive exposure to SARS-CoV-2, only one new asymptomatic infection occurred in this population. This may be due to unknown protective factors or chance. On the basis of this first descriptive study, using its population as a reference model, further investigations will be conducted to characterize the summer period exposure and to test the advanced hypotheses, focusing on the evaluation of a possible cross-reactivity to SARS-CoV-2 from exposure to endemic viruses.

## Introduction

Severe acute respiratory syndrome coronavirus 2 (SARS-CoV-2) is the causative agent of coronavirus disease (COVID-19).(1) It is a beta-coronavirus with a close structural and phylogenetic relationship to other pathogens of the “Coronaviridae” viral family, such as SARS-CoV-1(2) and MERS-CoV.(3)

While both the international and Italian scenarios were dominated by uncertainties, born out of the fragile balance between the need for drastic interventions and the inevitable economic repercussions, the virus found its way to “Isola del Giglio”, the same island off the coast of Tuscany, where, in 2012, the Italian cruise ship Costa Concordia ran aground and overturned after striking an underwater rock.

It remains one of the few places in the world where despite the arrival of the SARS-CoV-2, no case of symptomatic COVID19 has been diagnosed among the inhabitants. This observation has caught extremely broad international media coverage (4) and is the subject of this article.

## Methods

The aim of the study was to assess whether the observed “resistance” to SARS-CoV-2 had a serological foundation or to identify asymptomatic cases, seeking to detect SARS-CoV-2 antibodies through the screening process, simultaneously laying the foundation to future investigations focused on unraveling the causal dynamics of the apparent resistance.

The imported cases will be thoroughly described later in the article. All of these, except for one, reached the island prior to implementation of distancing measures, while being asymptomatic and later presented symptoms, exposing the population to significant infectious risk, as both phases have been associated with virus transmission.(5–9) Despite the prolonged exposure in a densely populated scenario, the infectious potential of the affected subjects (5–8) and the lack of distancing measures, the resident population did not develop COVID-19 syndromic features in the following months raising profound interest in the local medical community.

This study received ethical approval on April 25, 2020 by the Ethical Committee of the “Istituto Nazionale Malattie Infettive – Lazzaro Spallanzani”.

### Characterization of the exposure

#### (A) Geographic and demographic features of the island

Geographically, Giglio Island has a mountainous terrain, with ninety percent of its surface being inhabitable or inaccessible due to natural barriers. The last census dates back to May 2019, counting 1,423 inhabitants, in three main settlements. The island covers a surface of 24.1 km^2^ with a population density of 59 inhabitants/km.^2^

To estimate the island population density at the time of the serological screening, our study adjusted the surface area as illustrated in Table 1, excluding extra-urban lots, using the official structural plan and maps, provided as a guidance by the municipality and publicly available online.(10)

**Table 1.**
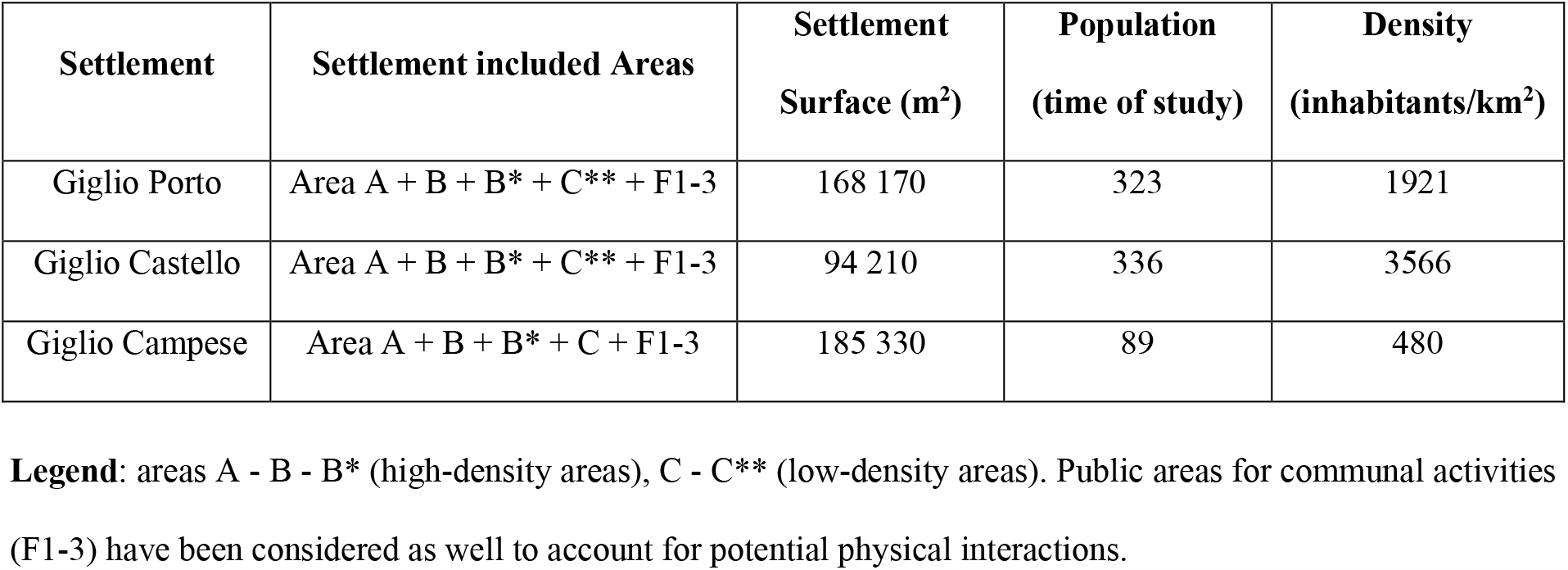
Geographic and Demographic characteristics of the Giglio Island.

| Settlement | Settlement included Areas | Settlement Surface (m<sup>2</sup>) | Population (time of study) | Density (inhabitants/km<sup>2</sup>) |
| --- | --- | --- | --- | --- |
| Giglio Porto | Area A + B + B* + C** + F1-3 | 168 170 | 323 | 1921 |
| Giglio Castello | Area A + B + B* + C** + F1-3 | 94 210 | 336 | 3566 |
| Giglio Campese | Area A + B + B* + C + F1-3 | 185 330 | 89 | 480 |

**Legend:** areas A - B - B\* (high-density areas), C - C\*\* (low-density areas). Public areas for communal activities (F1-3) have been considered as well to account for potential physical interactions.

Given the adjusted surface of 0.44771 km^2^, the resulting population density is 1670.7 inhabitants/km^2^. By omitting “Giglio Campese” to avoid a negatively skewed distribution, the resulting estimate in population density is 2511.62 pp/Km^2^, which would rank the local municipality as the 122^th^ most densely populated in Italy (Top 1.54 percentile out of 7903 Municipalities). This is an important information as population density is a likely factor correlated with spread of COVID-19.(11)

#### (B) Ferry connection between the island and the mainland

During lockdown, the connection to the mainland was through a ferry route with two daily trips covering 11 marine miles. Despite the maximum capacity of 600 passengers, the ferries carried a daily average of ten individuals, rendering the island practically isolated.

#### (C) Description of imported COVID-19 Cases

The data related to this section are available through direct contact with the corresponding author.

### Study Population

The population screening by serological rapid test on SARS-CoV-2 began on April 29 and ended on May 3, 2020. We initiated population screening for SARS-CoV-2 by inviting all adult residents who were present on the island. Eighty-five people, including healthcare professionals and volunteers, after testing negative at the serological antibody test, assisted in conducting the screening. The participation of island inhabitants was assessed and monitored through cross-checking the updated documents provided by the municipality.

The testing was performed in pre-determined locations. Subjects signed an informed consent. A health professional administered a questionnaire asking for background information (age,gender) permanence on the island and smoking status. All data were recorded in digital form. Each participant underwent rapid serological testing and provided a self-collected sample of saliva.

### Rapid Testing Serology Kit

The island population has been tested through the use of COVID-19 IgG/IgM Rapid Test Cassette (COVID-19 IgG/IgM Rapid Test Cassette (whole blood/serum/plasma), Product/Model: GCCOV-402a, Lot: 2003280, Zhejiang Orient Gene Biotech Co Ltd, Huzhou, Zhejiang, China). This rapid kit is a Lateral-Flow, solid-phase Immunochromatographic Assay (LFIA) for the rapid, qualitative and differential detection of IgG and IgM antibodies.

The studies conducted by the manufacturer, illustrated in Tables 3-4, report an overall test sensitivity (IgM or IgG) of 87.9% (87/99) and a specificity (IgM or IgG) of 100% (14/14). The sensitivity for solely IgG test was estimated to be 97.2% (35/36) during the convalescence period. An independent study, by Hoffman et al.(15) evaluated the same assay we used for the screening and reported a 69% sensitivity and 100% specificity for IgM detection, while denoting a 93.1% sensitivity and 99.2% specificity for IgG.

**Table 3.** Test overall (IgG or IgM) performance analysis as reported by Manufacturer.

| Method |  | RT-PCR |  | Total |
| --- | --- | --- | --- | --- |
|  |  | Positive | Negative |  |
| Covid-19 IgG/IgM Rapid<br>Test | Positive | 87 | 0 | 87 |
|  | Negative | 12 | 14 | 26 |
| Total |  | 99 | 14 | 113 |

**Table 4.** Test IgG sensitivity analysis as reported by Manufacturer.

| Method |  | COVID-19 patients in<br>convalescence | Total |
| --- | --- | --- | --- |
| Covid-19 IgG/IgM Rapid<br>Test | Positive | 35 | 35 |
|  | Negative | 1 | 1 |
| Total |  | 36 | 36 |

In manufacturer’s analyses the rapid kit’s specificity was comparable the results of both Hoffman’s study and the pooled results from a recent meta-analysis (MA) by Bastos et al.(16) On the other hand the analyses concerning the test sensitivity presented a much higher estimate when compared to the MA’s results on LFIAs’ sensitivity, pooled from 17 diagnostic test studies involving 1857 patients: 66.0% (95%CI: 49.3 to 79.3).

### Nucleic Acid Amplification Tests (NAAT) on saliva

Samples of saliva were refrigerated after collection and stocked in a facility on the island. Upon its completion, they have been transferred to the Onco-genomics and Epigenetics Unit of “IRCSS - Regina Elena National Cancer Institute” to avoid bias associated to point-of-care NAAT assays.(17) Viral genetic material was extracted using Nextractor® NX-48S DNA/RNA Extraction System (Genolution).

The saliva specimens have been analyzed with Allplex 2019-nCoV Assay (Seegene), a real-time reverse transcriptase polymerase chain reaction (RT-PCR) test.

The NAAT testing on saliva has been the object of mixed criticism with studies warning against a reduced sensitivity compared to the upper respiratory tract.(18,19) In contrast, other studies yielded a satisfactory sensitivity for NAATs self-collected saliva, suggesting that they might be a valid alternative to testing on nasopharyngeal samples, especially in settings with resources constraints.(20–23)

## Results

The study successfully screened a total of 723 people, including 634 of 748 residents present at the time of the study (84.8%), and 89 non-residents (including 85 healthcare professionals and Cases 3 and 4).

The screening population (residents and non-residents) consisted of 364 males and 359 females. Out of 723 individuals, 195 identified themselves as smokers, 175 as ex-smokers and 353 denied any previous direct smoking history. The currently smoking portion of the study population is largely made of cigarette-smokers (185 out of 195, 94.9%), with an average daily count of 14 cigarettes/day. The median age of the population was 58.5 years (IQR 27.9 years). The stratified analysis on the smoking population highlighted a slightly uneven gender distribution (108 males vs 87 females) with a median age of 48.7 years (IQR 22.4 years).

### Rapid Testing Serology Kit

The Rapid Testing Serology Kit identified only one previously undiagnosed positive asymptomatic subject by IgM positivity and confirmed the presence of anti-SARS-CoV-2 antibodies in the two tested patients previously recognized as positive by RT-PCR on the rhino-pharyngeal swab, for a total of 3 positives for anti-SARS-CoV-2 IgG/IgM out of 723 participants. The asymptomatic subject found IgM-positive was tested twice more to corroborate the finding. Case 1 could not be tested as deceased prior to study beginning; Cases 2 and 5 refused to be tested or provide a biological specimen.

### Nucleic Acid Amplification Tests on saliva

A total of 723 samples of saliva were collected from the population. Of these, 66 could not be analyzed due to collection of insufficient material. The NAAT on patients’ saliva did not identify any subject positive for SARS-CoV-2 RNA.

## Discussion

### Summary of findings

Despite of the introduction of SARS-CoV-2 on Giglio Island by five cases, known to have had physical interaction with local population, our study found one asymptomatic subject, positive to IgM but negative to RT-qPCR and confirmed the absence of either current or past viral infection in 633 residents of this highly populated area through qualitative assessment of anti-SARS-CoV-2 antibodies and RNA in saliva.

### Strengths and limitations

There are several strengths to our study. The fact that the island population remained isolated throughout the lockdown period creates a closed cohort model with few confounding factors and a reasonably well characterized exposure. Also, the data and sample collection of our study has been properly standardized on the basis of predetermined criteria, as to increase reproducibility and limit bias. The high participation rate also reduces concerns of selection bias.

There are also some limitations. A portion (15.2%) of the resident population did not participate to the serologic screening. This factor, associated to the inevitable lack of blinding among the islanders and the voluntary basis of participation, might have biased the results, underestimating the positive rate. The great variability in rapid serological test performances, particularly in regard of its sensitivity, might have led to overestimate the negative rate. The molecular tests, despite having a confirmatory role, given the time interval between the actual beginning of the study and the time of possible exposure, might have biased the results to finding no cases, failing to recognize a low viral count from the prior month’s infection. While the sample size is limited, it is naturally given by the island’s population. However, chance as a possible explanation of our findings cannot be excluded.

### Findings in comparison to other studies

There are few comparisons with other studies, because, despite sharing similarities with other seroprevalence investigations,(24) it refers to a small and quasi-segregated population where it was possible to carefully track every case of COVID-19 and the social network of exposed individuals related to it.

### Implications for research

Because at the time of arrival of four of the five documented COVID-19 cases, social distancing measures had not yet been put in practice and given the densely populated urban area, we inferred that the absence of confirmed cases is attributable to other factors, such as (A) external environmental determinants; (B) intrinsic determinants to the population; (C) the infectious agent; (D) chance.

Apart from chance, among the plausible environmental determinants, we hypothesize that the scarcity of air pollution might have played a role in limiting the COVID-19 spread. This hypothesis would be consistent with previous studies that highlighted a correlation between lack of air pollutants and low number of infections(25) and COVID-19 mortality.(26,27) Other hypothetical co-determinants are the peculiar geo-climatic and microenvironmental conditions present on the island, possibly reducing the viral load in the aerosol or limiting its infectiveness once exposure occurs.

Another theory to explain this apparent resistance of the population lies in the possibility of a cross-reactive immunity, conferring either protection or reducing the severity of COVID-19, after the exposure to a closely related pathogen. This hypothesis is consistent with some studies showing the presence of a SARS-CoV-2 reactive CD4 T cell subpopulation, manifesting cross reactivity with antigens of endemic coronaviruses, the causative agents of common cold.(28–30)

### Conclusions

Despite extensive exposure to SARS-CoV-2, only one new asymptomatic infection occurred in this population, as documented by IgM but not by RT-qPCR. This may be due to unknown protective factors or chance. On the basis of this first descriptive study, we will carry out further investigations to test the advanced hypotheses using its population as a reference model. In particular, our immediate research activities will focus on examining cross-reactivity to SARS-CoV-2, investigating a possible humoral contribution, both systemic (IgG/IgM mediated) and localized (IgA mediated), accounting also for potential interaction with the residing microbiota. We will also conduct a second serological screening following the population’s exposure during a busy summer season that brought thousands of tourists to the island. We will study whether and how the island inhabitants were potentially infected with SARS-Co V-2 during that period.

## Data Availability

The authors confirm that the data supporting the findings of this study are available within the article

## Acknowledgements

We would like to express our graditude to the population of Giglio Island who generously participated in the study, the island volunteers who directly worked on the study planning and implementation, including the “Confraternita Misericordia” in Giglio Island. We also thank the Municipality of Giglio Island, the Regional Health System local Unit “Toscana Sud-Est” Director and managers, the pharmacy in Giglio Island, the Association “Together in Pink” from Castiglione della Pescaia, the Grosseto General Hospital, the Carabinieri of Giglio island local station and Municipality Police for their important and efficient support to the study. Our graditude is also directed to Elisa Milano, Giulia Orlandi e Alina Catalina Palcau from the Oncogenomic and Epigenetic Unit at IRCCS Regina Elena National Cancer Institute, who all assisted in preparing the biologic sample for genetic testing. Furthermore, we thank the “Istituto Nazionale Malattie Infettive – Lazzaro Spallanzani”, and in particular both the Scientific Director Giuseppe Ippolito and the Institutional Review Board for their important inputs about the study implementation. Finally, we thank the General Director of the Tuscany Region for the strong support of the study and the availability to provide kits for serological assessment of the island population.

